# Infection by SARS-CoV-2 with alternate frequencies of mRNA vaccine boosting for patients undergoing antineoplastic treatment for cancer

**DOI:** 10.1101/2023.05.25.23290402

**Authors:** Jeffrey P. Townsend, Hayley B. Hassler, Brinda Emu, Alex Dornburg

**Affiliations:** Department of Biostatistics, Yale School of Public Health, New Haven, Connecticut 06510, USA; Program in Computational Biology and Bioinformatics, Yale University, New Haven, Connecticut 06511, USA; Department of Ecology and Evolutionary Biology, Yale University, New Haven, Connecticut 06525, USA; Interdisciplinary Graduate Program in Quantitative Biosciences, Georgia Institute of Technology, Atlanta, GA 30332, USA; Department of Internal Medicine (Infectious Diseases), Yale University, New Haven, Connecticut 06437, USA; Department of Bioinformatics and Genomics, University of North Carolina, Charlotte, North Carolina 28223, USA

## Abstract

Patients undergoing antineoplastic therapies often exhibit reduced immune response to COVID-19 vaccination, necessitating assessment of alternate boosting frequencies for these patients. However, data on reinfection risks to guide clinical decision-making is limited. We quantified reinfection risks of SARS-CoV-2 at different mRNA boosting frequencies of patients on antineoplastic therapies. Antibody levels following Pfizer-BioNTech BNT162b2 vaccination were analyzed for patients without cancer, with cancer undergoing various treatments, and treated with different antineoplastic therapeutics. Using long-term antibody data from other coronaviruses in an evolutionary framework, we estimated infection probabilities based on antibody levels and projected waning. We calculated cumulative probabilities of breakthrough infection for alternate booster schedules over two years. Annual boosting reduced risks for targeted or hormonal treatments, immunotherapy, and chemotherapy-immunotherapy combinations similarly to the general population. Patients receiving no treatment or chemotherapy exhibited higher risks, suggesting that accelerated vaccination schedules should be considered. Patients treated with rituximab therapy posed the highest infection risk, suggesting that a combination of frequent boosting and additional interventions may be warranted for mitigating SARS-CoV-2 infection in these patients.

## Background

Compared to individuals in the general population, many patients undergoing diverse antineoplastic therapies exhibit a reduced immune response to COVID-19 vaccination. Therefore, potential benefits of alternate frequencies of boosting for these patients should be assessed.

## Objective

Quantify reinfection risks of SARS-CoV-2 under alternate frequencies of mRNA boosting for patients undergoing common antineoplastic therapies.

## Methods and Findings

We obtained published antibody (anti-RBD) levels following Pfizer-BioNTech BNT162b2 vaccination of patients without cancer, with cancer undergoing no treatment, or treated with various antineoplastic therapeutics—targeted or hormonal, Hematopoietic Stem Cell Transplantation (HSCT), immunotherapies, chemotherapy, immunotherapy & chemotherapy, or rituximab (1). To project antibody waning based on initial responses for each patient cohort in ongoing therapy, we applied an established comparative evolutionary framework (2–4) for inference of infection probability given antibody level to mRNA boosting with the BNT162b2 vaccine. We integrated longitudinal anti-N and anti-S IgG antibody waning data for six human-infecting coronaviruses (HCoV-OC43, HCoV-NL63, and HCoV-229E, SARS-CoV-1, SARS-CoV-2, and MERS; 3) into an ancestral and descendent states analysis, fitting logistic regression models of endemic daily probabilities of infection without additional interventions (2,4). From ensuing daily probabilities of infection given antibody level, cumulative probabilities of breakthrough infection were calculated for alternate, variant-updated booster schedules for members of the general population, patients with untreated cancer and undergoing continuous treatment over two years, scheduled every month, three months, six months, one year, or two years.

For patients undergoing targeted or hormonal treatments, immunotherapy, a combination of chemotherapy and immunotherapy, or HSCT therapy, annual boosting led to a 2.5-fold reduction in risk over two years relative to foregoing boosters (12–14% vs 29–31%), a result similar to that expected for non-cancer patients. For patients undergoing chemotherapy alone, risks were higher: 18% breakthrough for those with annual boosting, 8% with boosting every six months, and 3% with boosting every three months. Notably, for cancer patients receiving no current treatment, the risks of breakthrough infection were worse than for any of these treatments: over two years, infection was projected to occur at rates of 22%, 11%, and 6% for yearly, six-month, or three-month booster schedules. Notably, this level of benefit was not achievable for patients undergoing rituximab therapy, for which the probability of breakthrough infection reached 18% even with boosting monthly. Nearly 2 out of 5 patients were predicted to experience breakthrough infections with annual boosting.

**Figure.**
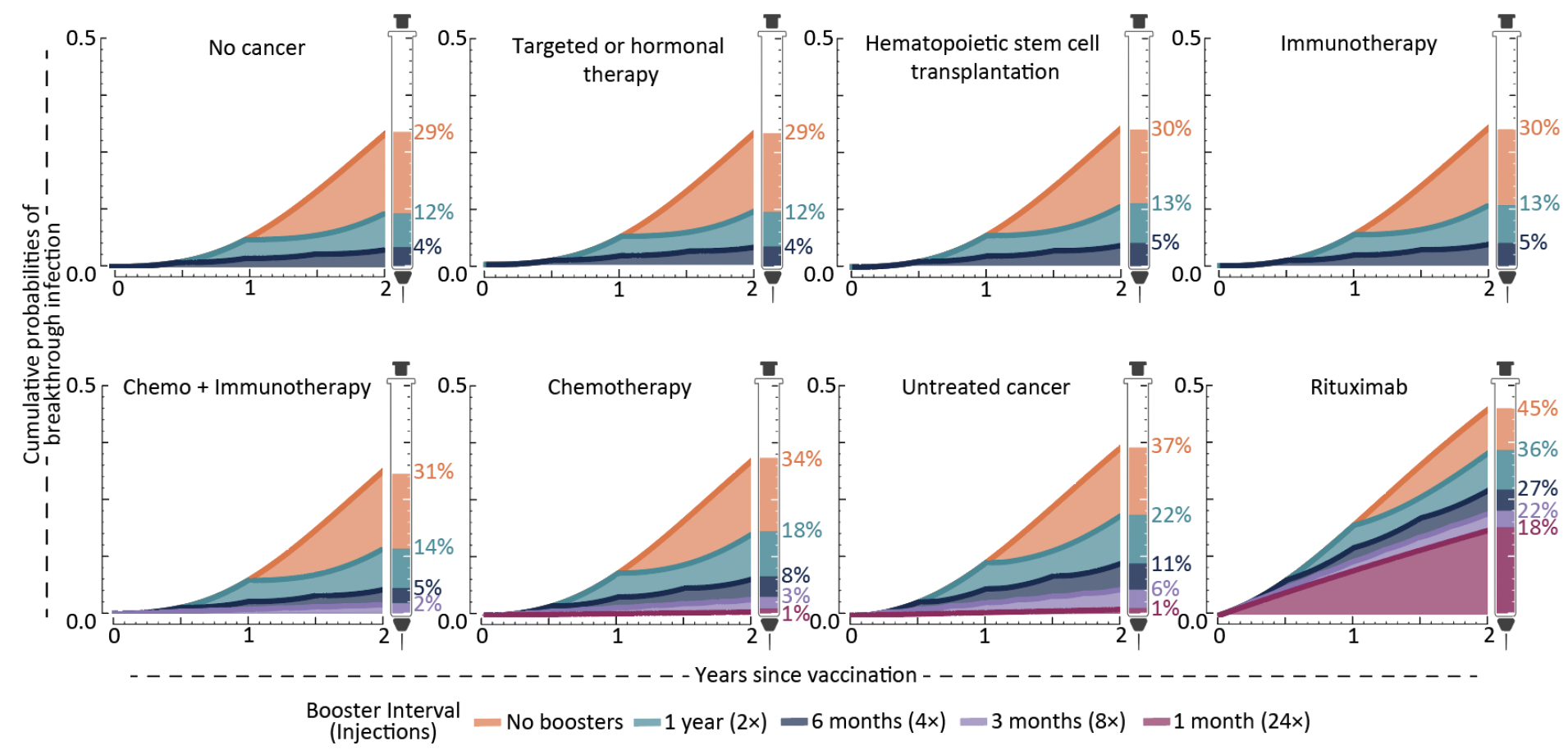
Cumulative probabilities of breakthrough infection with alternate frequencies of updated BNT162b2 booster vaccination following the primary series, for the general population, for patients undergoing six antineoplastic therapies (targeted or hormonal therapy, hematopoietic stem cell transplantation, immunotherapy, chemotherapy plus immunotherapy, chemotherapy, and rituximab), and for patients with untreated cancer.

## Discussion

Here we have employed antibody levels in response to vaccination, their waning rates, and endemic daily probabilities of infection based on antibody levels to quantify the probability of breakthrough SARS-CoV2 infection using alternate booster schedules for patients undergoing antineoplastic therapies. For patients undergoing targeted or hormonal therapy, HSCT, immunotherapy, and the combination of immunotherapy and chemotherapy, we found that benefits of regular boosting largely match those for the general population: increased protection with increased frequency of boosting. For patients undergoing chemotherapy alone or with untreated cancers, breakthrough infection risks are elevated, and merit consideration for an accelerated vaccination schedule. Moreover, treatment with the B-cell depleting monoclonal antibody rituximab—prescribed for some hematologic malignancies and also as an immune-modulating treatment for other diseases such as rheumatoid arthritis—poses substantially higher short- and long-term risk of SARS-CoV-2 infection regardless of booster strategy. Indeed, our finding of a higher risk of infection parallels previous findings that patients treated with rituximab are at higher risk for severe COVID-19 symptoms (5). Therefore, supplementary interventions such as masking, isolation, and use of prophylactic antibodies targeting SARS-CoV-2 are warranted.

Booster vaccines for SARS-CoV-2 are being developed to target predominant, emerging strains. Our study incorporates this updating as well as the post-uptake waning of vaccine efficacy due to antigenic evolution. However, our quantification of infection risks does not incorporate the component of antigenic evolution that occurs during the *circa* three-month delay between manufacturing and deployment of mRNA booster vaccinations. This delay can reduce booster efficacy. However, over these short timescales, vaccines generally remain efficacious. Our assessment provides knowledge for clinical decision-making that can have substantial impact on the mitigation of potentially severe SARS-CoV-2 infections in cancer patients undergoing antineoplastic therapies. Further research incorporating antibody responses to vaccines provided during other treatments would facilitate increasingly comprehensive estimation of the benefits of specific schedules of booster vaccination that mitigate risks for vulnerable subsets of the population.

## Data Availability

Reproducible Research Statement: All data, code, and a detailed readme have been archived on Zenodo: DOI:10.5281/zenodo.7963444.

https://zenodo.org/record/7963444

